# Multiparametric Quantitative MRI of Peripheral Nerves to Differentiate Demyelinating from Axonal Polyneuropathies

**DOI:** 10.1101/2025.11.21.25340765

**Authors:** Jesus E. Fajardo, Vivian B. Truong, Yang Xuan, Sara E. Benitez, Mary L. Vo, Bo Hu, Richard D. Dortch, Jun Li, Yongsheng Chen

**Affiliations:** Department of Neurology, Wayne State University School of Medicine, Detroit, MI, USA; Department of Neurology, Houston Methodist Research Institute, Houston, TX, USA; Department of Translational Neuroscience, Barrow Neurological Institute, Phoenix, AZ, USA

**Author notes:** Correspondence to Dr. Yongsheng Chen Dr. Jun Li. J. Li and Y. Chen are joint senior authors. **Acknowledgement** We gratefully acknowledge the participants and the MR technologists at Wayne State University and Houston Methodist Research Institute. The authors thank Stephanie Yan Xuan for her assistance in study coordination. **Author Contributions JEF**: writing – original draft, formal analysis, visualization. **VBT**: data curation, project administration. **YX**: data curation, project administration. **SEB**: data curation, investigation. **MLV**: data curation, investigation, validation. **BH**: data curation, investigation, resources. **RDD**: methodology, resources. **JL**: conceptualization, resources, supervision, funding acquisition. **YC**: conceptualization, resources, supervision, funding acquisition, writing – original draft. All authors reviewed and revised the final draft. **Competing Interests** None declared.

## Abstract

**Background:** Differentiating demyelinating from axonal polyneuropathies is essential for accurate diagnosis and treatment. We hypothesized that multiparametric quantitative MRI (qMRI) of peripheral nerves can differentiate demyelination from axonal loss. This retrospective study leveraged genetically defined demyelinating and axonal polyneuropathies to test this concept.

**Methods:** Multiparametric qMRI data of proximal (sciatic) and distal (tibial) nerves were acquired on 3T MRI, including magnetization transfer ratio (MTR), MT saturation index (MTsat), T *, T, proton density (PD), fractional anisotropy (FA), mean/axial/radial diffusivities (MD, AD, RD), and fascicular volume (fVol). Data were analyzed from patients with Charcot-Marie-Tooth type 1 (CMT1, de-/dys-myelinating, n=19), CMT2 (axonal, n=12), hereditary neuropathy with liability to pressure palsies (HNPP, a cohort who often has intermediate changes between the two classifications, n=25), and health controls (HC, n=25). A composite qMRI score, as CMT Imaging Score (CMTIS), was developed to predict disease severity using the CMT Neuropathy Score version-2 (CMTNSv2) as a clinical reference. Receiver operating characteristic (ROC) analyses assessed diagnostic performance.

**Results:** CMT1 showed significantly increased fVol versus HCs, while CMT2 demonstrated reduced T_2_*. Both CMT1 and CMT2 exhibited reduced FA, MTsat, and AD, along with elevated T_1_ and RD, with larger abnormalities in CMT1. ROC analyses demonstrated strong discrimination of CMT1 and CMT2 (AUCs: 0.95 and 0.85 for sciatic; 0.89 and 0.73 for tibial nerves). CMTIS correlated strongly with CMTNSv2 (r=0.67 sciatic; r=0.72 tibial; r=0.79 combined).

**Conclusions:** Multiparametric qMRI identifies distinct imaging signatures of demyelinating versus axonal hereditary polyneuropathies. The CMTIS shows strong potential as a biomarker for disease monitoring.

**DATA AVAILABILITY:** Anonymized data used in this study is available from the corresponding author upon request and subject to institutional approvals.

**KEY MESSAGES:** *What is already known on this topic:* Current electrophysiological tools are limited in their ability to differentiate demyelinating from axonal polyneuropathies when pathology occurs in proximal nerves. Quantitative MRI (qMRI) can assess proximal demyelination and axonal loss; however, individual qMRI metrics lack sufficient sensitivity for reliable differentiation.

*What this study adds:* This proof-of-concept study demonstrates the feasibility of using multiparametric qMRI for patient stratification and for distinguishing demyelinating from axonal inherited polyneuropathies. The proposed composite qMRI score shows a strong correlation with clinical disease severity.

*How this study might affect research, practice, or policy:* This study suggests that multiparametric qMRI of peripheral nerves can serve as a non-invasive adjunct to distal nerve conduction studies for improving diagnosis and treatment management of polyneuropathies. The composite qMRI score also shows potential as a monitoring biomarker for tracking disease progression.

## INTRODUCTION

Polyneuropathies are a diverse group of disorders affecting peripheral nerves, typically involving demyelination or axonal degeneration/loss ^1–3^, with the latter resulting to long-term disability ^4^.

Demyelination may induce secondary axonal loss and conduction block, causing acute deficits. These impairments also denervate skeletal muscle, leading to intramuscular fat accumulation ^5^.

Nerve conduction studies (NCS) are a standard tool for diagnosing demyelinating and axonal neuropathies. The former typically shows slowed conduction velocities, conduction block, and/or temporal dispersion, while the latter is characterized by reduced amplitudes in compound muscle action potentials and sensory nerve action potentials ^6–9^. However, NCS is largely limited to assessing distal nerve segments and often becomes non-responsive when most nerve fibers are damaged (i.e., in length-dependent neuropathies) ^5^. If demyelination occurs in the proximal nerves, NCS cannot access the proximal pathology, even though late responses may suggest proximal conduction block, but not specific ^10^. Accurately diagnosing demyelinating neuropathies is important for guiding treatment decisions. For inherited neuropathies, genetic mutations causing demyelinating versus axonal polyneuropathies are distinct. With the emergence of gene therapies, the two groups of polyneuropathies require different outcome measurements and therapeutic strategies. For acquired polyneuropathies, chronic inflammatory demyelinating polyneuropathy (CIDP) responds well to immunomodulatory therapies such as intravenous immunoglobulin, glucocorticoids, or plasma exchange, but axonal polyneuropathies do not respond to these treatments ^11, 12^. Thus, differentiating between the two types affects clinical decisions. There remains a significant gap in non-invasive, reliable biomarkers for differentiating demyelinating from axonal types for patients with lesions in proximal nerves that are not reliably accessible by distal NCS.

This gap can be addressed with quantitative magnetic resonance imaging (qMRI) of peripheral nerves, a non-invasive technique capable of assessing both proximal and distal nerves using various qMRI parameters sensitive to demyelination and axonal loss ^13^. For instance, relaxation qMRI parameters such as T_1_, proton density (PD), and T_2_* are sensitive to myelin and water content changes ^13^; magnetization transfer ratio (MTR) ^14^ and MT saturation index (MTsat) ^15^ provide myelin specific information; and fractional anisotropy (FA), mean diffusivity (MD), axial diffusivity (AD), and radial diffusivity (RD) derived from diffusion tensor imaging (DTI) are sensitive to tissue microstructural integrity ^16–18^. However, given the challenges in imaging the small target of peripheral nerves, single qMRI parameter may not reliably assess nerve pathologies ^13^. To this end, multiparametric qMRI protocols have been developed to simultaneously quantify these measures and provide complementary information for a more reliable assessment ^19^. We hypothesize that multiparametric qMRI of peripheral nerves can predict probabilities of demyelination and axonal loss for individual patients with polyneuropathies. The multiparametric qMRI of proximal and distal nerves can serve as a supplement to distal NCS in diagnosis and treatment management.

The objectives of this study are twofold. First, we test the hypothesis by using multiparametric qMRI data from patients with genetically confirmed demyelinating and axonal subtypes. Second, we explore the potential of a composite qMRI score derived from multiparametric qMRI metrics as an imaging biomarker of disease severity. Data from health controls (HC) and three patient cohorts with Charcot–Marie–Tooth (CMT) diseases were included in this retrospective study: a demyelinating cohort of CMT type 1 (CMT1), an axonal cohort of CMT type 2 (CMT2), and a cohort with hereditary neuropathy with liability to pressure palsies (HNPP), which often shows intermediate features between these two classifications ^1,20^.

## MATERIALS AND METHODS

### Standard Protocol Approvals, Registrations, and Patient Consents

The study was approved by the local institutional review board (IRB) at Wayne State University (WSU, Detroit, MI, USA) and Houston Methodist Hospital (HMH, Houston, TX, USA) following NIH’s single IRB requirement (protocol #020519MP2F). Written consent was acquired for each participant at enrollment.

### Participants and Clinical Assessment

Participants were enrolled from WSU and HMH between 2021 to 2025. The inclusion criteria were set as male or female participants from 18 to 90 years old who have been diagnosed with polyneuropathy, such as CMT, CIDP, HNPP, or idiopathic polyneuropathy. The exclusion criteria included: 1) inability to fit in the knee coil, 2) MRI contraindications, 3) pregnancy, or 4) orthopedic implants on both lower limbs. Volunteers without a history of neuropathy were also enrolled as HCs. All patients were seen by the physician (J.L.) for clinical assessment. The CMT

Examination Score (CMTES) was obtained by neurologists, and NCS of the forearms and legs were performed by experienced technicians as described in publications ^21–23^. Subsequently, the CMT Neuropathy Score v2 (CMTNSv2) ^24^ was calculated, reflecting overall neurological disabilities. CMTES was calculated by using physical findings and excluding NCS readouts.

Finally, available data from HC and genetically confirmed CMT1, CMT2, and HNPP were used for this retrospective study.

### MRI Data Acquisition and Processing

MRI scans were performed on each participant’s right leg using 3.0-T MRI scanners with a knee coil (Siemens Verio and Cima.X at WSU; Siemens Vida at HMH). A standardized qMRI protocol was used to image the sciatic nerve (Sn) at the mid-thigh and tibial nerve (Tn) at the mid-calf levels ^19^. Briefly, the two-session qMRI protocol employed a set of vendor-supplied 3D gradient echo (GRE) sequences to compute nerve fascicular volume (fVol) in a resolution of 0.15 × 0.15 × 3.0 mm^3^, MTR, MTsat, T_2_*, T_1_, PD in a resolution of 0.6 × 0.6 × 3.0 mm^3^; and a DTI sequence in a resolution of 1.2 × 1.2 × 6.0 mm^3^ to compute FA, MD, AD, and RD metrics. There were 40 and 16 axial slices for the GRE and DTI sequences, respectively. The qMRI protocol was about 30 minutes per location. Detailed imaging parameters are shown in Table S1 of the supplementary material.

Regions-of-interest (ROIs) for the Sn and Tn fascicles were segmented using an established semi-automated method on the fat-suppressed GRE images as previously described ^13, 19^. Nerve fVol was computed by multiplying voxel count by voxel size. Mean values within each ROI were extracted for all qMRI parameters using the validated pipeline in MATLAB, as previously described ^13–15, 19, 25–27^. PD values were normalized to the adjacent normal-appearing muscle signal. All results were exported to spreadsheets for statistical analysis.

### Descriptive Statistics

All statistical analyses were performed using custom Python scripts developed with Scikit-learn ^28^, Numpy ^29^ and SciPy ^30^. Data from all subjects are reported as mean ± standard deviation, along with the 5th to 95th percentile range. Two-sided t-tests were carried out for each qMRI parameter to assess the statistical significance of the differences between cohorts (CMT1, CMT2, HNPP and HC). A one-way ANOVA was conducted on the qMRI data across phenotypes to assess its separability. Statistical significance level was set as p < 0.05. Since the comparison of individual qMRI parameters between the patient and HC cohorts was intended to identify multiparametric qMRI patterns and select a subset of parameters for the separation model, multiple-comparison corrections were not applied to the two-sided t-tests. In contrast, significance levels from the ANOVA tests were reported using Bonferroni corrections.

### Polyneuropathy Classification using qMRI

To test the hypothesis that multiparametric qMRI of peripheral nerves can differentiate demyelinating from axonal polyneuropathies, receiver operating characteristic (ROC) analysis with logistic regression (LR) models were performed for the qMRI data of the Sn and Tn from the 4 cohorts: CMT1 as the demyelinating group, CMT2 as the axonal group, HNPP as a group with mixed demyelination and axonal loss, and HC as the control group. All features were standardized using z-score normalization to ensure comparability across variables before the LR model classification. A subset of parameters with F-score higher than 10 were selected from Sn and Tn, including fVol, MTsat, T_1_, T *, PD, FA, and RD. This threshold was selected to ensure stronger discriminatory power between groups, based on established practices in medical imaging literature, where F-scores below 10 are usually excluded due to weak class separation ^31,32^. A stratified (80% train - 20% validation) 5-fold cross-validation strategy was employed to ensure balanced representation of classification in each fold. For each fold, the model was trained on the training set and evaluated on the validation set. L2 regularization was applied to reduce the overfitting likelihood. To evaluate the model’s ability to distinguish specific classes, ROC curves were plotted, and the area under the ROC curve (AUC) was calculated. For results robustness, ROC curves were averaged across folds, and the mean true positive rate (TPR) was interpolated over a common false positive rate (FPR) grid. Mean AUC values were reported for each of the 3 LR models – qMRI of Sn, qMRI of Tn, and qMRI of both nerves.

Finally, the 3 LR models were verified on a temporal validation dataset unseen by the models to confirm their performance in individual patient classification. The testing dataset was from available one-year follow-up scans on 24 participants.

### CMT Imaging Score to Predict Disease Severity

To derive the CMT Imaging Score (CMTIS), a composite qMRI score predictive of CMTNS and CMTES, a linear regression model was trained using Python Scikit-learn ^28^. Stratified 5-fold cross-validation was employed to preserve the distribution of polyneuropathy subgroups (CMT1, CMT2, HNPP) across folds. For each training fold, a linear regression model was fit, and the average coefficient of all linear regressions was used as final coefficient for each qMRI parameter. Details of the CMTIS calculation are shown in the supplementary material.

## RESULTS

### Demographics and clinical data

There were 80 participants’ qMRI data for the sciatic nerve (CMT1 = 19, CMT2 = 12, HNPP = 24, and HC = 25), and 80 subjects for the tibial nerve (CMT1 = 19, CMT2 = 12, HNPP = 25, and HC = 24). Demographic characteristics, including age, sex, body mass index (BMI), and number of patients with clinical scores (CMTES and CMTNSv2), are summarized in **Table 1**.

**Table 1.**
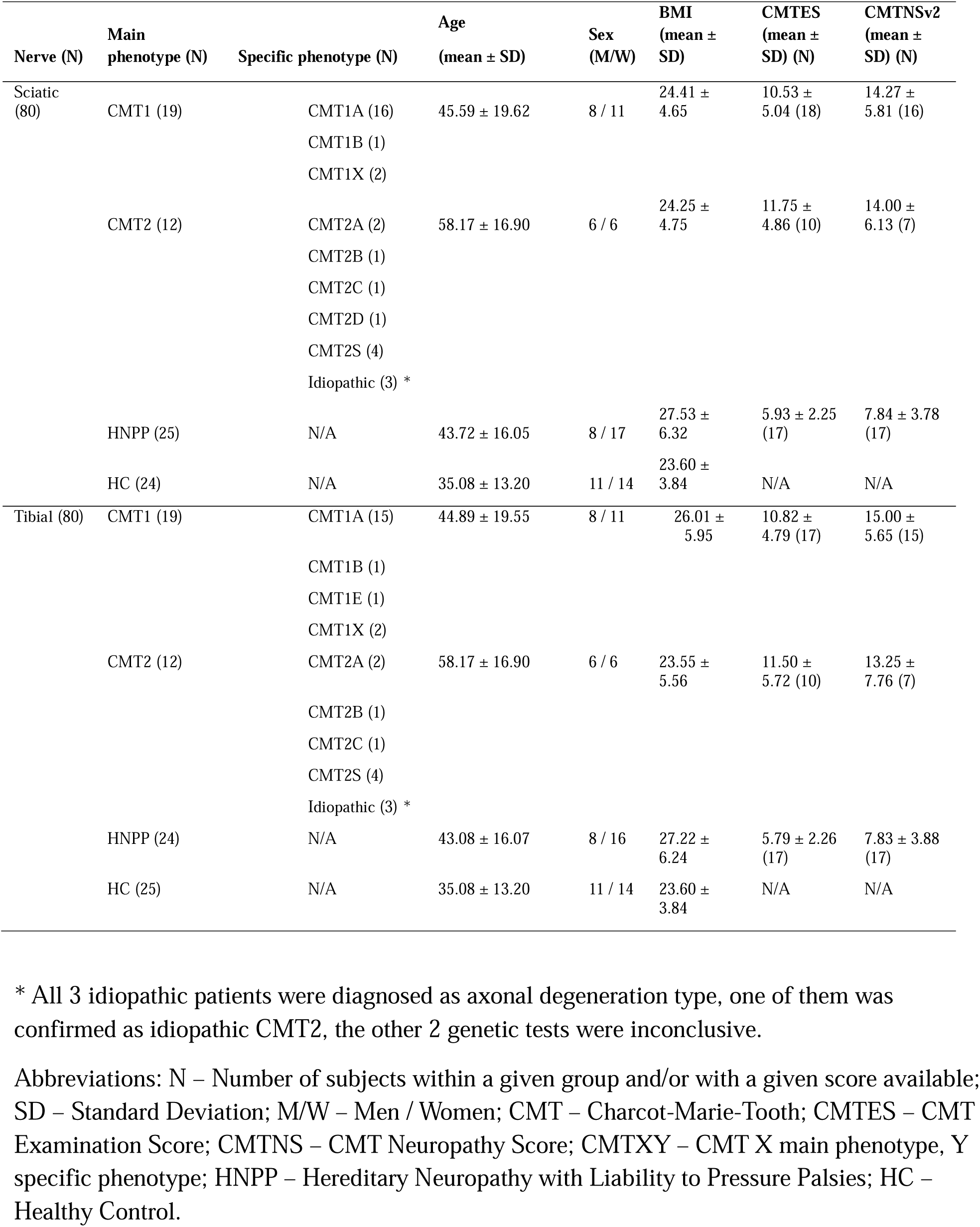
Demographics. * All 3 idiopathic patients were diagnosed as axonal degeneration type, one of them was confirmed as idiopathic CMT2, the other 2 genetic tests were inconclusive. Abbreviations: N – Number of subjects within a given group and/or with a given score available; SD – Standard Deviation; M/W – Men / Women; CMT – Charcot-Marie-Tooth; CMTES – CMT Examination Score; CMTNS – CMT Neuropathy Score; CMTXY – CMT X main phenotype, Y specific phenotype; HNPP – Hereditary Neuropathy with Liability to Pressure Palsies; HC – Healthy Control.

Representative images of Sn and Tn across cohorts are shown in **Figure 1**.

**Figure 1.**
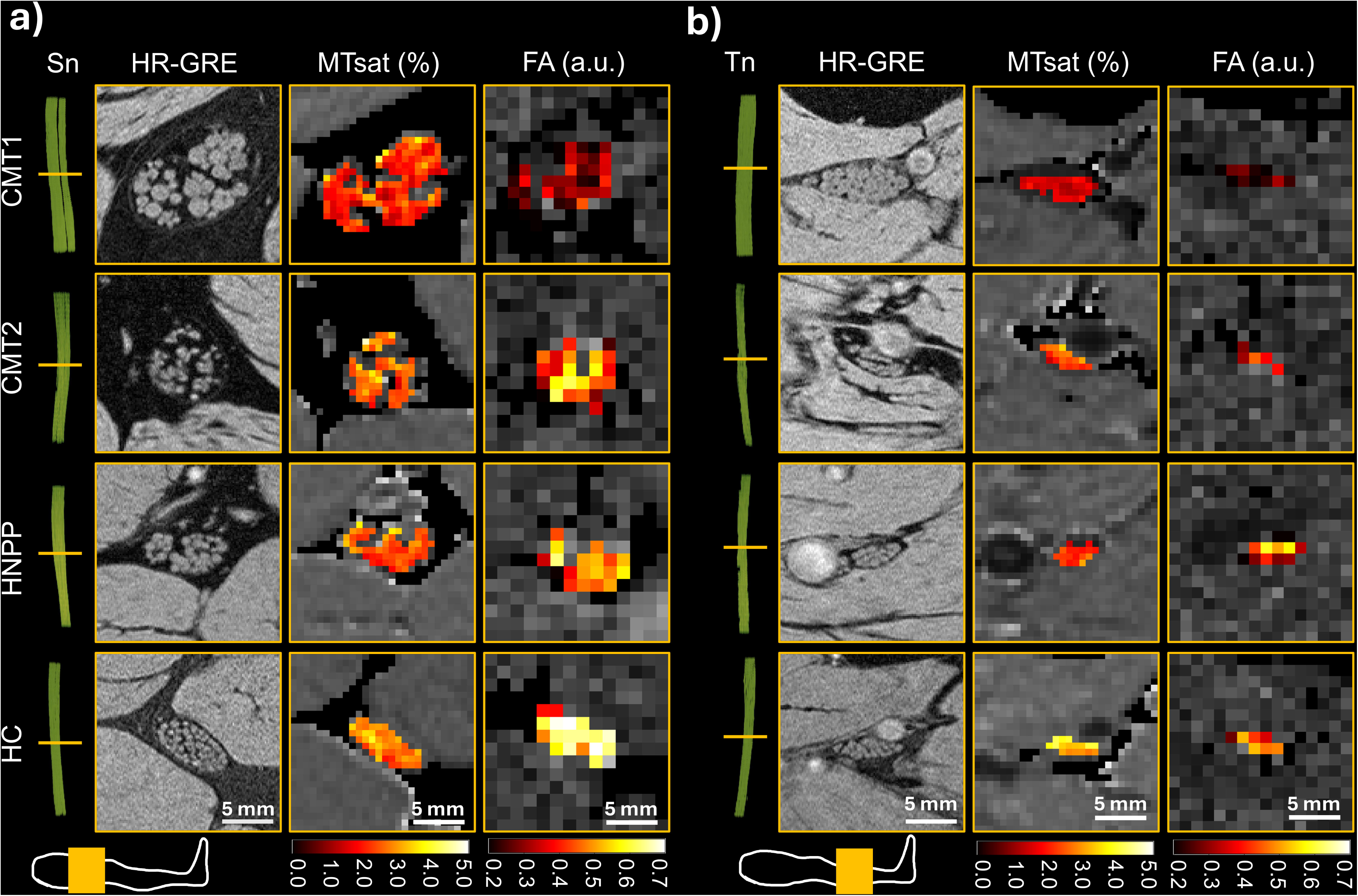
Representative multiparametric qMRI images from each participant cohort. Images of the Sn and Tn are shown in the panel a) and b), respectively. In the first column, a 3D rendering of the nerve length is shown with a yellow bar indicating the images cross-section. The leg scheme in each panel indicates the coil position in yellow. Subsequently, images from the high-resolution (HR) GRE with a resolution of 0.15 × 0.15 × 3.0 mm^3^, MTsat maps with a resolution of 0.6 × 0.6 × 3.0 mm^3^, and FA maps with a resolution of 1.2 × 1.2 × 6.0 mm^3^ are shown for the CMT1, CMT2, HNPP, and HC participants. The colored pixels were the pixels in the ROI for extracting the qMRI value for the participant. The resolution and ROIs for MTR, T_2_*, T_1_, and PD maps were the same as those for the MTsat. Similar, MD, AD, and RD were using the resolution and ROI the same as those for the FA map.

### Multiparametric qMRI Patterns of Sciatic and Tibial Nerves in Different Polyneuropathies

As shown in **Figure 2**, qMRI of the Sn for CMT1 patients exhibited the most pronounced alterations, including significantly reduced MTR, MTsat, FA and AD, and elevated T_1_, PD, MD and RD, compared to HC. Nerve size measured by fVol was exclusively increased in CMT1, consistent with nerve hypertrophy. In contrast, CMT2 patients showed a reduced PD, but more moderate reductions in MTsat, FA and AD, and elevated T_1_ and RD. The fVol and MD in CMT2 were comparable with those in HC. There was a significantly lower T_2_* in CMT2. In contrast, the T_2_* was unremarkable in CMT1 and HNPP. HNPP patients demonstrated intermediate changes, with significant reductions in MTsat, FA and AD, and elevations in T_1_, PD and RD, though less severe than in CMT1. Patterns of changes with respect to HC for the evaluated qMRI parameters are summarized in **Table 2** to facilitate the differentiation between demyelinating and axonal types.

**Figure 2.**
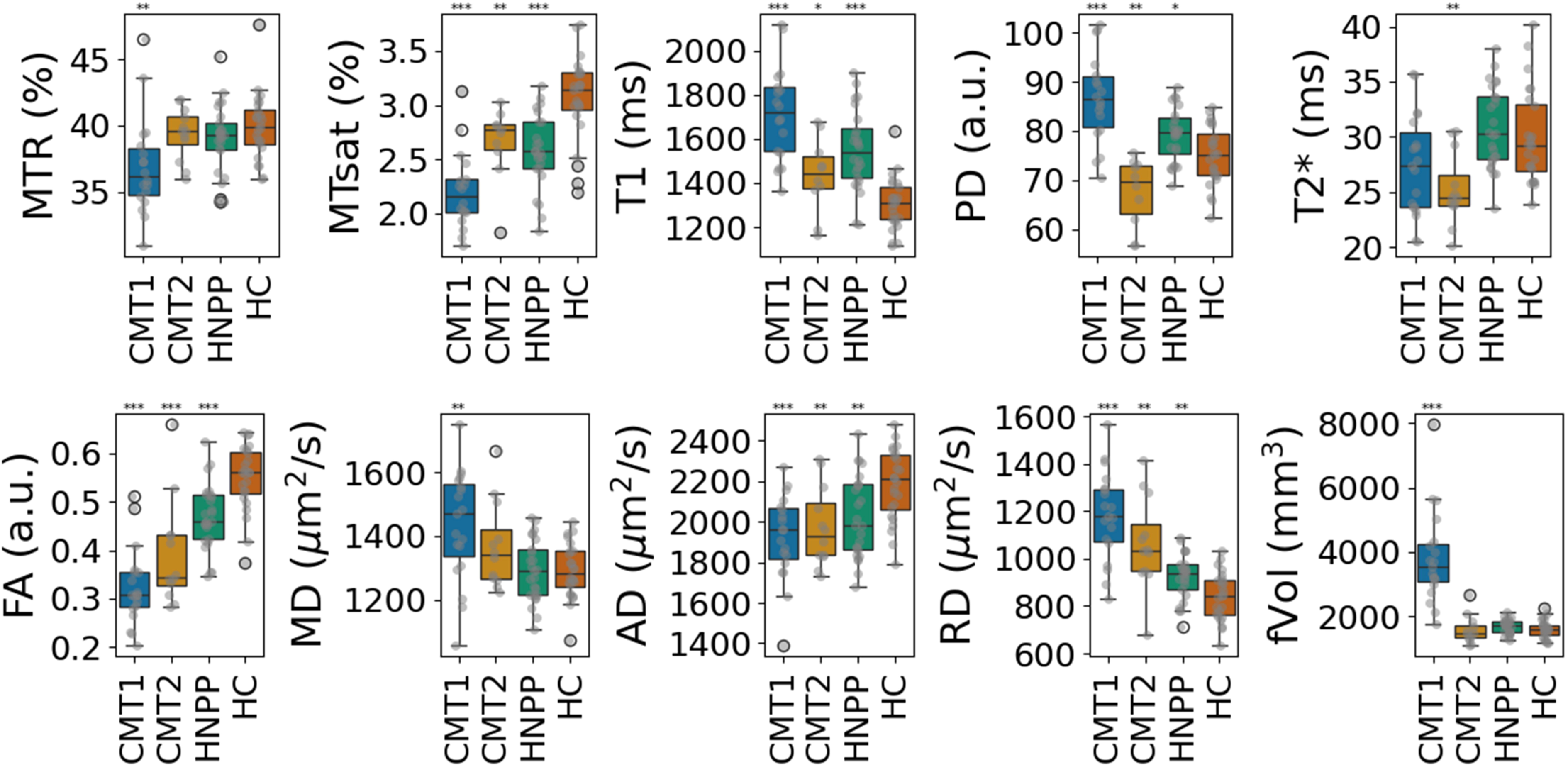
Multiparametric qMRI of sciatic nerve. Box plots with values of MTR, MTsat, T_1_, PD, T_2_*, FA, MD, AD, RD and fVol for CMT1 (blue boxes), CMT2 (mustard boxes), HNPP (green boxes) patients and HC (orange boxes). Data points are shown in gray over the box plots. Stars are shown over the corresponding polyneuropathy group, indicating the level of statistical significance of that group compared to HC: * p < 0.05; ** p < 0.001; *** p <0.0001.

**Table 2.** Upward and downward trends with respect to HC for the multiparametric qMRI parameters at the Sn and Tn levels.

| **Polyneuropathy** | **Sciatic Nerve** |  |
| --- | --- | --- |
|  | **Parameters with elevated values** | **Parameters with decreased values** |
| CMT1 | T<sub>1</sub>, PD, MD, RD, fVol | MTR, MTsat, FA, AD |
| CMT2 | T<sub>1</sub>, RD | MTsat, FA, AD, PD, T<sub>2</sub><sup>*</sup> |
| HNPP | T<sub>1</sub>, PD, RD | MTsat, FA, AD |
|  | **Tibial Nerve** |  |
|  | **Parameters with elevated values** | **Parameters with decreased values** |
| CMT1 | T<sub>1</sub>, PD, MD, RD, fVol | MTR, MTsat, FA, AD |
| CMT2 | T<sub>1</sub>, RD | MTsat, FA, AD, T<sub>2</sub><sup>*</sup> |
| HNPP | T<sub>1</sub>, PD, MD, RD | MTsat, FA, AD |

Abbreviations: qMRI – Quantitative MRI; CMT – Charcot-Marie-Tooth; CMTX – CMT X main phenotype; HNPP – Hereditary Neuropathy with Liability to Pressure Palsies; HC – Healthy Control; T<sub>1</sub> – Longitudinal magnetic relaxation time; PD – Proton Density; T<sub>2</sub><sup>\*</sup> - Transverse magnetic relaxation including local magnetic field inhomogeneities effect; MTR – Magnetization Transfer Ratio; MTsat – Magnetization Transfer Saturation; FA – Fractional Anisotropy; MD – Mean Diffusivity; AD – Axial Diffusivity; RD – Radial Diffusivity; fVol – Fascicles Volume.

Similar trends were observed for the Tn data (**Figure 3**). MTsat, FA and AD were significantly reduced in all patient groups compared to HC, with the most substantial reductions in CMT1. T_1_, PD, MD and RD were elevated in CMT1, HNPP and CMT2, however PD and MD didn’t reach statistical significance for CMT2, with more modest changes. T * was significantly reduced in CMT2. Nerve size measured by fVol was again significantly increased in CMT1 but remained unchanged in CMT2 and HNPP compared to HC. The values presented as boxplots in **Figure 2** and **Figure 3** are listed for reference in **Table 3**.

**Figure 3.**
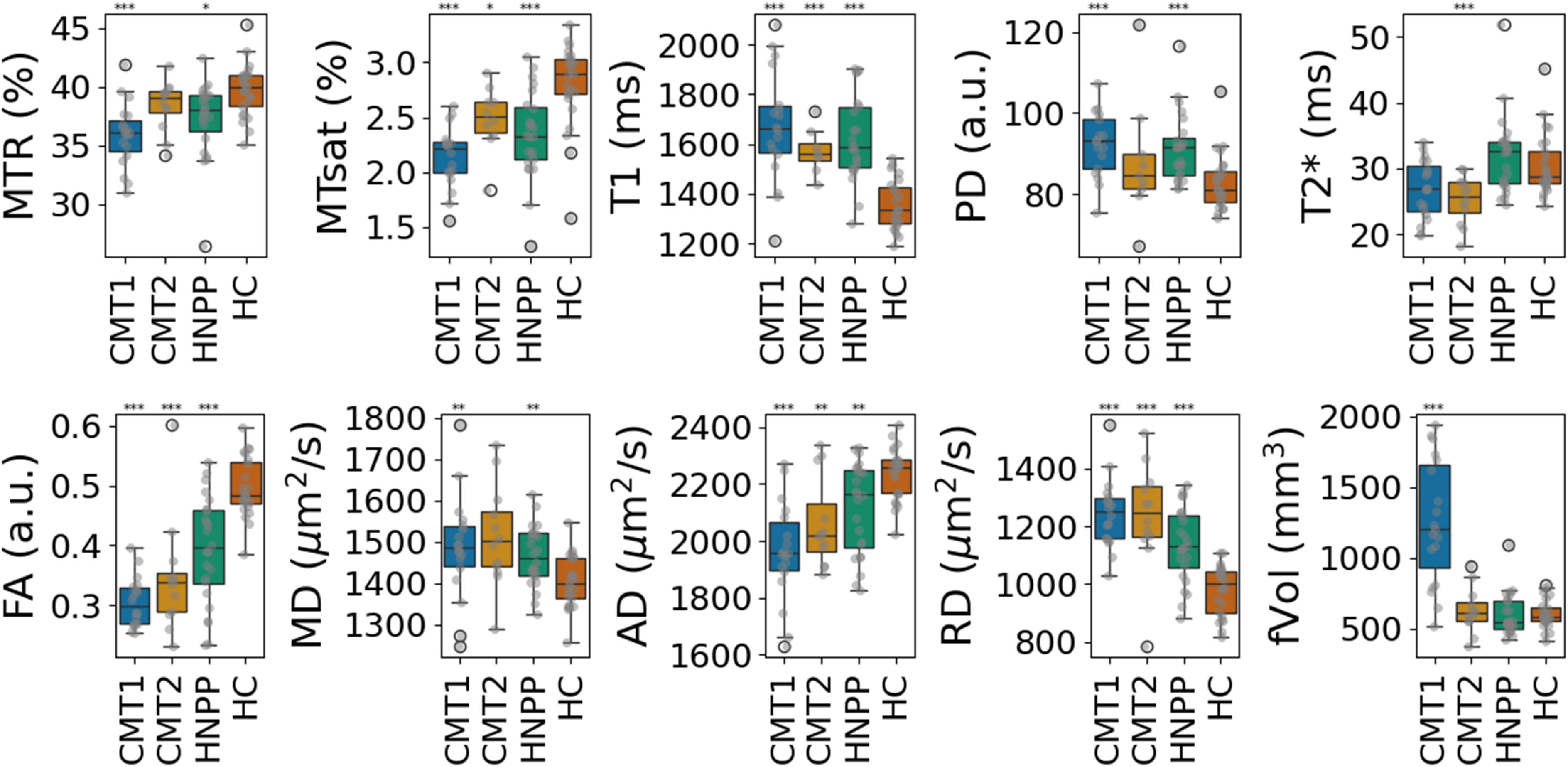
Multiparametric qMRI of tibial nerve. Box plots with values of MTR, MTsat, T_1_, PD, T_2_*, FA, MD, AD, RD and fVol for CMT1, CMT2 and HNPP patients and HC, are displayed in the same format of Figure 2.

**Table 3.** Multiparametric qMRI values for cohorts of CMT1, CMT2, HNPP, and HC in the Sn and Tn.

| qMRI Parameter | HC<br>Mean ± SD<br>(5%-95% percentile) | HNPP<br>Mean ± SD<br>(5%-95% percentile) | CMT1<br>Mean ± SD<br>(5%-95% percentile) | CMT2<br>Mean ± SD<br>(5%-95% percentile) |
| --- | --- | --- | --- | --- |
| **Sciatic nerve (Sn)** |  |  |  |  |
| T<sub>1</sub> (ms) | 1305.40 ± 120.72<br>(1125.80 - 1462.00) | 1550.12 ± 183.88<br>(1125.80 - 1462.00) *** | 1689.06 ± 196.94<br>(1436.90 - 1935.26) *** | 1435.07 ± 171.65<br>(1174.95 - 1670.65) * |
| PD (a.u.) | 75.02 ± 6.03 (66.00 - 83.80) | 79.25 ± 5.46 (72.20 - 86.96) * | 85.77 ± 8.47 (73.03 - 100.39) *** | 67.64 ± 6.95 (56.80 - 74.85) ** |
| T<sub>2</sub><sup>*</sup> (ms) | 29.97 ± 4.28 (25.67 - 37.96) | 30.84 ± 3.71 (26.60 - 36.24) | 26.91 ± 4.29 (20.58 - 32.76) * | 25.24 ± 3.27 (20.95 - 30.46) ** |
| MTR (%) | 39.95 ± 2.53 (36.22 - 42.68) | 38.99 ± 2.68 (34.32 - 42.40) | 37.18 ± 3.69 (33.12 - 44.16) * | 39.38 ± 1.97 (36.23 - 41.93) |
| MTsat (%) | 3.07 ± 0.39 (2.31 - 3.68) | 2.57 ± 0.34 (1.99 - 3.06) *** | 2.24 ± 0.35 (1.82 - 2.84) *** | 2.66 ± 0.34 (2.09 - 2.98) ** |
| FA (a.u.) | 0.54 ± 0.06 (0.42 - 0.64) | 0.46 ± 0.06 (0.35 - 0.57) *** | 0.32 ± 0.08 (0.22 - 0.49) *** | 0.39 ± 0.11 (0.28 - 0.58) *** |
| MD ($\mu\text{m}^2/\text{s}$) | 1292.50 ± 88.44<br>(1191.10 - 1414.10) | 1291.79 ± 96.78<br>(1151.07 - 1443.57) | 1430.11 ± 176.61<br>(1161.49 - 1624.97) ** | 1369.26 ± 135.92<br>(1233.46 - 1593.19) |
| AD ($\mu\text{m}^2/\text{s}$) | 2183.6 ± 178.65<br>(1898.70 - 2417.80) | 2024.68 ± 203.91<br>(1758.78 - 2304.10) ** | 1925.18 ± 216.56<br>(1595.43 - 2192.87) *** | 1972.49 ± 197.42<br>(1744.99 - 2298.94) ** |
| RD ($\mu\text{m}^2/\text{s}$) | 843.29 ± 101.06<br>(706.65 - 995.47) | 924.25 ± 96.60 (778.82 - 1071.25) ** | 1177.22 ± 192.89<br>(883.381 - 1445.02) *** | 1065.92 ± 196.64<br>(819.58 - 1356.24) ** |
| fVol (mm<sup>3</sup>) | 1614.40 ± 286.32<br>(1184.89 - 2069.32) | 1681.64 ± 228.07<br>(1335.62 - 2009.70) | 3733.66 ± 1426.85<br>(2071.71 - 6012.57) *** | 1588.62 ± 443.12<br>(1117.92 - 2357.72) |
| **Tibial nerve (Tn)** |  |  |  |  |
| T<sub>1</sub> (ms) | 1353.58 ± 101.00<br>(1228.91 - 1515.89) | 1622.68 ± 174.78<br>(1355.86 - 1897.30) *** | 1673.48 ± 222.67<br>(1363.82 - 2005.37) *** | 1571.35 ± 100.31<br>(1457.81 - 1705.31) *** |
| PD (a.u.) | 83.12 ± 6.87 (75.40 - 91.87) | 91.47 ± 8.41 (81.98 - 104.02) *** | 92.33 ± 8.01 (80.96 - 102.36) *** | 88.22 ± 15.17 (72.25 - 112.62) |
| T<sub>2</sub><sup>*</sup> (ms) | 30.55 ± 4.68 (25.64 - 37.89) | 31.93 ± 5.91 (25.36 - 40.20) | 26.85 ± 4.44 (20.78 - 33.26) * | 25.31 ± 3.63 (19.21 - 29.92) *** |
| MTR (%) | 39.71 ± 2.20 (36.40 - 42.79) | 37.32 ± 3.26 (33.78 - 40.17) * | 36.16 ± 2.66 (32.00 - 40.08) *** | 38.42 ± 2.16 (34.71 - 40.81) |
| MTsat (%) | 2.80 ± 0.38 (2.21 - 3.20) | 2.35 ± 0.39 (1.74 - 2.95) *** | 2.14 ± 0.25 (1.75 - 2.50) *** | 2.49 ± 0.30 (2.05 - 2.85) * |
| FA (a.u.) | 0.49 ± 0.048 (0.43 - 0.56) | 0.39 ± 0.08 (0.24 - 0.51) *** | 0.30 ± 0.04 (0.25 - 0.37) *** | 0.34 ± 0.11 (0.24 - 0.53) * |
| MD ($\mu\text{m}^2/\text{s}$) | 1409.56 ± 67.12<br>(1339.188 - 1526.83) | 1466.98 ± 72.77<br>(1355.120 - 1580.09) *** | 1506.60 ± 112.54<br>(1338.77 - 1685.41) *** | 1466.99 ± 124.44<br>(1355.10 - 1850.30) *** |
| AD ($\mu\text{m}^2/\text{s}$) | 2236.55 ± 91.87<br>(2110.28 - 2366.33) | 2117.36 ± 161.24<br>(1851.87 - 2321.02) ** | 1999.21 ± 182.96<br>(1722.45 - 2280.78) *** | 2062.31 ± 158.75<br>(1897.00 - 2317.34) ** |
| RD ($\mu\text{m}^2/\text{s}$) | 980.07 ± 87.24<br>(837.92 - 1101.09) | 1136.32 ± 129.29<br>(930.46 - 1318.81) *** | 1256.37 ± 113.31<br>(1120.16 - 1435.96) *** | 1237.65 ± 185.82<br>(972.44 - 1477.03) *** |
| fVol (mm<sup>3</sup>) | 602.50 ± 99.81<br>(463.46 - 787.09) | 598.49 ± 149.76 (451.85 - 770.08) | 1247.20 ± 423.73<br>(618.91 - 1881.84) *** | 602.50 ± 99.81<br>(463.46 - 787.09) |

Abbreviations: qMRI – Quantitative MRI; SD – Standard Deviation; CMT – Charcot-Marie-Tooth; CMTES – CMT Examination Score; CMTNS – CMT Neuropathy Score; CMTX – CMT X main phenotype; HNPP – Hereditary Neuropathy with Liability to Pressure Palsies; HC – Healthy Control; T<sub>1</sub> – Longitudinal magnetic relaxation time; PD – Proton Density; T<sub>2</sub><sup>\*</sup> – Transverse magnetic relaxation including local magnetic field inhomogeneities effect; MTR – Magnetization Transfer Ratio; MTsat – Magnetization Transfer Saturation; FA – Fractional Anisotropy; MD – Mean Diffusivity; AD – Axial Diffusivity; RD – Radial Diffusivity; fVol – Fascicles Volume.

ANOVA tests confirmed significant separability of polyneuropathy types based on multiparametric qMRI metrics (Table S2). The strongest discriminators (F-statistic > 10) were MTsat, T_1_, PD, FA, RD, and fVol in both the Sn and Tn.

### Polyneuropathy Classification using Logistic Regression

Logistic regression models trained on selected qMRI parameters (T_1_, T_2_*, PD, MTsat, FA, RD, fVol) demonstrated high diagnostic performance (**Figure 4**). Classification of CMT1 yielded AUC values of 0.95 (Sn), 0.89 (Tn), and 0.95 (combined data of Sn and Tn). CMT2 classification yielded AUCs of 0.85 (Sn), 0.73 (Tn), and 0.79 (combined data of Sn and Tn).

**Figure 4.**
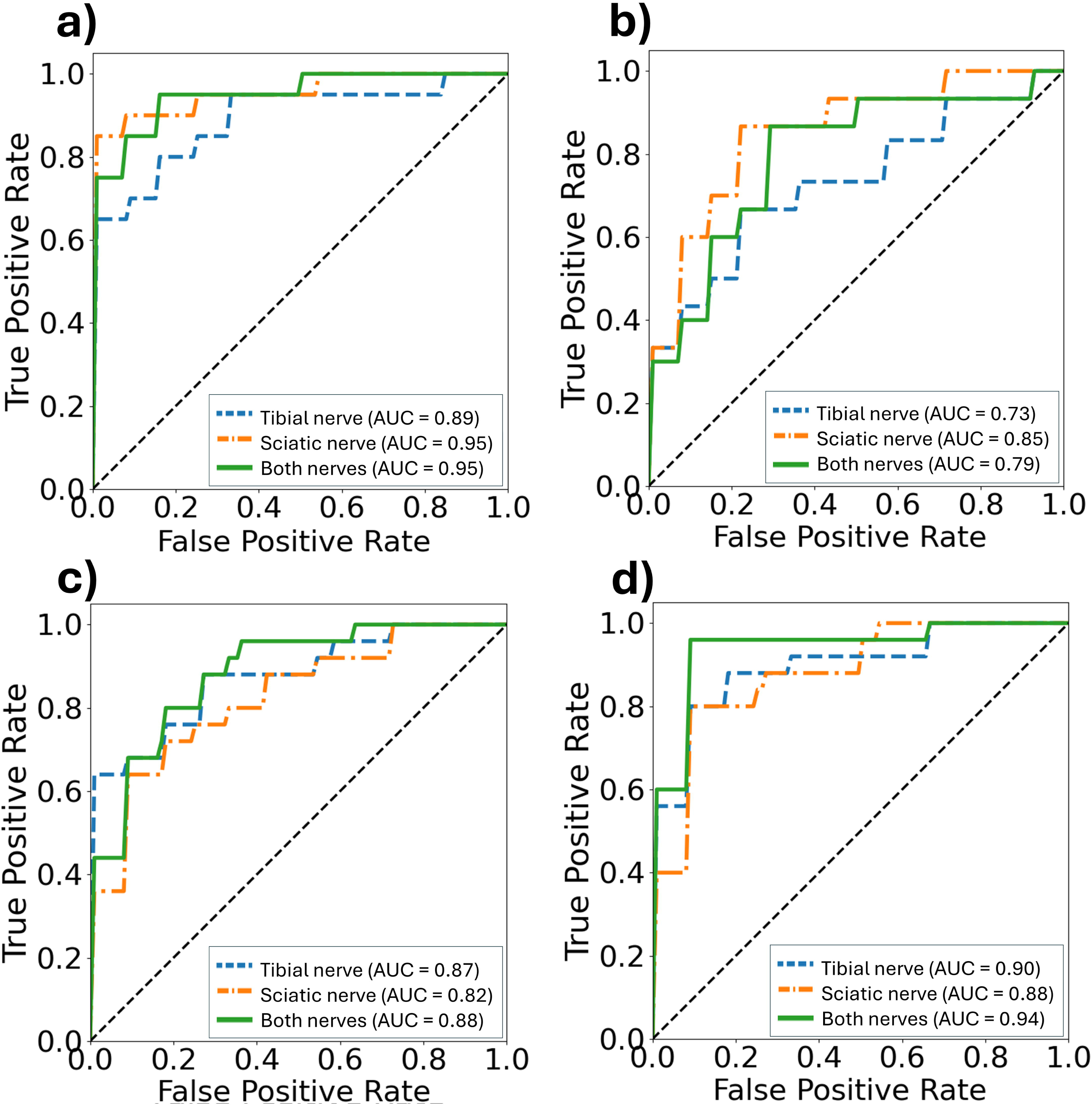
ROC analyses for subtype classification. ROC curves (True positive rate vs. False positive rate) from a logistic regression model are displayed using the following combination of selected parameters: T_1_, T_2_*, PD, MTsat, FA, RD, fVol. For predicting a) CMT1, b) CMT2, c) HNPP and d) HC. In dashed blue lines, the model results are shown for the Tn data, in dotted-dashed orange lines for the Sn data and in solid green using both Sn and Tn data. In the lower-right corner, the AUC values are shown for each curve.

Classification of HNPP and HC also showed robust performance (AUC > 0.82). There was no additional benefit in model performance observed by combining Sn and Tn data.

Results from the temporal testing dataset, comprising data not previously seen by the model, are shown in Figure S3 as a confusion matrix. Predicted probabilities of demyelination and axonal loss for individuals are presented in Table S5 with similar prediction performance to that reported in **Figure 4**.

### CMTIS as a Potential Biomarker of Disease Severity

The normalized CMTIS derived from linear regression of imaging parameters showed a significant correlation with clinical scores (**Figure 5**). For CMTES, correlation coefficients were r = 0.66 (Sn), r = 0.80 (Tn), and r = 0.88 (combined). For CMTNSv2, correlation coefficients were r = 0.67 (Sn), r = 0.72 (Tn), and r = 0.79 (combined), respectively. Variance decomposition (see Table S3) revealed that diffusion metrics (FA, MD, RD, AD) and T_2_* contributed most to the CMTIS, especially in the Sn.

**Figure 5.**
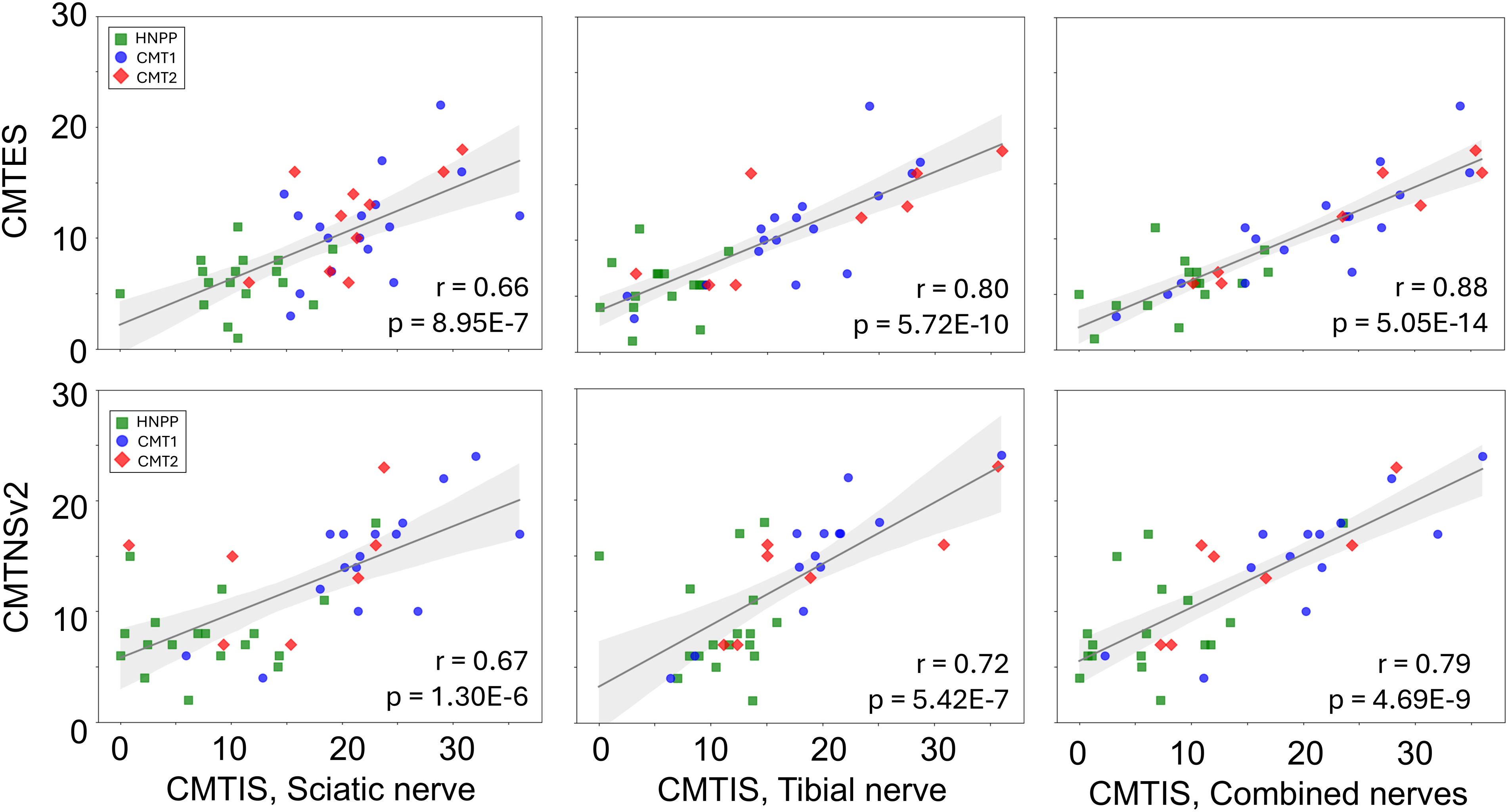
Correlations between CMTES/CMTNSv2 and the CMTIS. In the first row, scatterplots of CMTES vs. CMTIS (linear combination of T_2_*, FA, MD and RD) are shown alongside the result of a linear regression (gray line) showing in light gray the 5%-95% confidence interval of the regression. Green squares correspond to HNPP patients, blue dots correspond to CMT1 patients, and red diamonds to CMT2 patients. The correlation coefficient (r) and p-values are shown for each plot. In the second row, scatterplots of CMTNSv2 vs. CMTIS (linear combination of T_2_*, FA, RD, AD) are shown in the same format of the first row.

In Figure S1 and S2, the correlations of all individual qMRI parameters with CMTES and CMTNS in the Sn and Tn are displayed. Individual parameters with moderate-to-high correlation with CMTES in the Sn are T_2_* (r =-0.49; p = 0.0006), FA (r =-0.45; p = 0.002), RD (r = 0.43; p = 0.003) and with CMTNS are FA (r =-0.58; p = 0.0001), RD (r = 0.44; p = 0.003) and fVol (r = 0.52; p = 0.0005), consistent with the variance decomposition results of Table S3. Similarly in the Tn, T * (r =-0.40; p = 0.007) and fVol (r = 0.43; p = 0.003) have moderate-to-high correlation with CMTES and only fVol (r = 0.53; p = 0.0005) with CMTNS.

## DISCUSSION

While NCS is widely used to diagnose and evaluate nerve pathophysiological changes, it remains limited in assessing proximal nerves and is often uninformative on severely degenerated distal nerves. We hypothesize that multiparametric qMRI of peripheral nerves could complement NCS by improving the differentiation between demyelination and axonal loss in individual patients. In this study, we analyzed qMRI data from the sciatic and tibial nerves of patients with genetically confirmed inherited polyneuropathies to test whether qMRI can distinguish demyelinating from axonal polyneuropathies. Our findings revealed distinct qMRI signatures for demyelinating (CMT1), and axonal (CMT2) patients. ROC analyses showed strong predictive ability for demyelination and axonal loss for individual patients. Furthermore, the CMTIS had a strong correlation with disability scores measured by the CMTES and the CMTNSv2. These findings support the CMTIS as a potential objective monitoring biomarker in longitudinal studies to track disease progression. Longer-term, this approach may provide vital information for therapeutic decision-making when extended to acquired polyneuropathies.

### Polyneuropathy Classification by Multiparametric qMRI

The increased classification power of multiparametric qMRI compared to single qMRI parameters is demonstrated by the higher AUC observed when using a logistic regression model with an optimized combination of qMRI parameters, as compared to using single parameters (see Table S4). More data are needed to clarify whether the lower performance for identifying CMT2 (0.73 < AUC < 0.85) compared to CMT1 (0.89 < AUC < 0.95) relates to secondary axonal loss in CMT1 or the smaller sample size of the current CMT2 cohort. For the CMT1 and CMT2 groups, the sciatic nerve qMRI alone provides better model performance, likely due to the larger size of the proximal nerve, which reduces partial volume effects and underscores the value of proximal nerve qMRI for more reliable assessment where pathologies are not accessible by NCS. It should be noted that the trends in **Figure 4** changed slightly when the cross-validation seed changed; however, the AUC values remained relatively stable when running the model with 5 different seeds, yielding standard deviations of less than 5%. Outliers misclassified from the testing set (See Figure S3 and Table S5 of the Supplementary Material) were mainly due to overlapping distributions.

### CMTIS as a Potential Monitoring Biomarker for Disease Progression

The estimated CMTIS showed a strong and significant correlation with CMTES and CMTNSv2 scores, which suggests the qMRI has the potential to assess disease severity and may be used to monitor disease progression longitudinally. Importantly, the CMTIS is objective and avoids the subjectivity and inter-rater variability inherent in clinical scales, making it a potentially more reliable and reproducible tool. The distal nerve CMTIS showed a stronger correlation with neuropathy scores than the CMTIS of the proximal nerve, consistent with more severe pathologies in distal nerves, a length-dependent process in patients with inherited polyneuropathies. As shown in Table S3, these correlations were largely dominated by diffusion parameters (FA, MD, AD, and RD) and T *. Combining the Sn and Tn qMRI data further increased the correlation with the CMTES and CMTNSv2 scores. The fVol parameter was deliberately excluded from the CMTIS, due to the lack of correlation between this parameter and polyneuropathies other than CMT1 (see Figures S1 and S2), consistent with what has been previously reported in the literature ^2, 3^.

### Microstructural Implications in Multiparametric qMRI

Multiparametric qMRI revealed distinct patterns of pathologies between CMT1 (demyelinating) and CMT2 (axonal) which is consistent with the established pathological differences between the two conditions. CMT1 patients exhibited significantly reduced MTsat and FA, alongside elevated T_1_, PD, RD, and fVol, which is consistent with widespread demyelination (or dysmyelination), increased water content, and nerve hypertrophy ^14, 33^. The strong reduction in FA, driven primarily by increased RD rather than AD, suggests that transverse diffusivity is a key marker of myelin disruption in this group, consistent with previous studies ^16, 17^.

MTR and MTsat are both sensitive to myelin content, although they exhibit subtle but important differences sensitivity to confounding features. More specifically, the enhanced performance of MTsat is likely due to its reduced dependence on T_1_ relaxation and B_1_ field inhomogeneities, which can confound MTR measurements ^15^. By incorporating corrections for these confounding factors, MTsat provided a more specific and quantitative assessment of myelin integrity (MTR vs MTsat ANOVA F-statistic: 4.09 vs. 19.52 (Sn) and 9.03 vs. 16.27 (Tn), respectively), making it a potentially more promising biomarker for detecting demyelination in polyneuropathies.

To address slab-profile inhomogeneity effects (see *Methods* section), oversampling was increased along the partition dimension. This adjustment had minimal impact on qMRI normative values in healthy controls, aside from an approximately 10% increase in T_1_ compared with our previous work ^19^.This correction updates the normative T_1_ values for the Sn and Tn and provides a more accurate reference for interpreting pathological changes.

Although the MRI data were acquired on three different 3T scanners (Siemens Verio, Vida, and Cima.X), our harmonized acquisition protocols and standardized offline post-processing pipeline used to compute qMRI parameters helped to ensure cross-scanner validity. Specifically, MTR, MTsat, T_1_, and PD were derived from multiple scans, which inherently minimizes sequence-specific signal variations arising from data reconstruction and scanner calibration. In addition, B_1_ corrections were performed within the post-processing pipeline to further minimize hardware dependent signal variations ^19, 25^. R * was computed from multi-echo acquisitions within a single scan, further reducing systematic variability across scanners. Finally, DTI parameters depend primarily on the processing algorithm, which was standardized in the offline pipeline^27^ rather than relying on scanner-generated diffusivity maps. That said, real-world inter-scanner validation remains an important next step to ensure broader applicability.

### Limitations

Several limitations should be considered when interpreting these findings. First, the cohorts were not age-matched; in particular, the health controls were younger than those CMT patients.

Although nerve qMRI may exhibit physiological changes with age, these are generally much smaller than the pathological changes in polyneuropathies ^19, 34, 35^. Second, the present study was limited to common inherited polyneuropathies and did not include patients with acquired polyneuropathies. This will be the focus of future studies, as a tool that can differentiate demyelinating from axonal pathology is likely to have substantial impact in this patient population. Third, although the correlations between the CMTIS and clinical scores (CMTNSv2 and CMTES) were statistically robust, these results should be interpreted with caution due to the relatively small sample size (n ≈ 40). Future large-cohort prospective studies involving both acquired and inherited polyneuropathies with longitudinal designs are needed to determine the reliability of multiparametric qMRI, its sensitivity to disease progression, its correlation with clinical outcomes, and its potential to serve as an objective monitoring biomarker for both disease progression and therapeutic response.

## CONCLUSION

In this study, we demonstrated that multiparametric qMRI can differentiate de/dysmyelinating from axonal forms of inherited polyneuropathy at both the Sn and Tn levels. The qMRI metrics revealed distinct patterns across these polyneuropathy types. These findings justify future efforts in differentiating acquired demyelinating from axonal polyneuropathies, with the goal of developing a deployable diagnostic tool alongside current clinical standard. Additionally, the CMTIS derived from these qMRI parameters showed promise as an objective imaging biomarker of disease severity, exhibiting strong correlations with clinical disability scores and supporting its potential use for monitoring progression in future clinical trials. These findings support the utility of standardized qMRI protocols in the non-invasive assessment and monitoring of polyneuropathies.

## Data Availability

Anonymized data used in this study is available from the corresponding author upon request and subject to institutional approvals.

## REFERENCES

1. Li J. Inherited neuropathies. Seminars in neurology; 2012: Thieme Medical Publishers: 204–214.

2. Pušnik L, Lechner L, Serša I, et al. 3D fascicular reconstruction of median and ulnar nerve: Initial experience and comparison between high-resolution ultrasound and MR microscopy. European Radiology Experimental 2024;8:100.

3. Shibuya K, Sugiyama A, Ito Si, et al. Reconstruction magnetic resonance neurography in chronic inflammatory demyelinating polyneuropathy. Annals of neurology 2015;77:333–337.

4. Park SB, Cetinkaya-Fisgin A, Argyriou AA, Höke A, Cavaletti G, Alberti P. Axonal degeneration in chemotherapy-induced peripheral neurotoxicity: clinical and experimental evidence. Journal of Neurology, Neurosurgery & Psychiatry 2023;94:962–972.

5. Li J, Ghandour K, Radovanovic D, et al. Stoichiometric alteration of PMP22 protein determines the phenotype of hereditary neuropathy with liability to pressure palsies. Archives of neurology 2007;64:974–978.

6. Pareyson D, Marchesi C. Diagnosis, natural history, and management of Charcot–Marie–Tooth disease. The Lancet Neurology 2009;8:654–667.

7. Daube JR, Rubin DI. Nerve conduction studies. Aminoff’s Electrodiagnosis in Clinical Neurology [Internet] 6th ed Philadelphia, PA 2012:289–325.

8. Lewis RA, Sumner AJ, Shy ME. Electrophysiological features of inherited demyelinating neuropathies: a reappraisal in the era of molecular diagnosis. Muscle & nerve 2000;23:1472–1487.

9. Zifko UA, Zipko HT, Bolton CF. Clinical and electrophysiological findings in critical illness polyneuropathy. Journal of the neurological sciences 1998;159:186–193.

10. Kiers L, Clouston P, Zuniga G, Cros D. Quantitative studies of F responses in Guillain-Barré syndrome and chronic inflammatory demyelinating polyneuropathy. Electroencephalography and Clinical Neurophysiology/Evoked Potentials Section 1994;93:255–264.

11. Mair D, Madi H, Eftimov F, Lunn MP, Keddie S. Novel therapies in CIDP. Journal of Neurology, Neurosurgery & Psychiatry 2025;96:38–46.

12. McCray BA, Scherer SS. Axonal Charcot-Marie-Tooth disease: from common pathogenic mechanisms to emerging treatment opportunities. Neurotherapeutics 2021;18:2269–2285.

13. Chen Y, Haacke EM, Li J. Peripheral nerve magnetic resonance imaging. F1000Res 2019;8:1803.

14. Dortch RD, Dethrage LM, Gore JC, Smith SA, Li J. Proximal nerve magnetization transfer MRI relates to disability in Charcot-Marie-Tooth diseases. Neurology 2014;83:1545–1553.

15. Helms G, Dathe H, Kallenberg K, Dechent P. High resolution maps of magnetization transfer with inherent correction for RF inhomogeneity and T1 relaxation obtained from 3D FLASH MRI. Magnetic Resonance in Medicine: An Official Journal of the International Society for Magnetic Resonance in Medicine 2008;60:1396–1407.

16. Chhabra A, Carrino JA, Farahani SJ, et al. Whole body MR neurography: Prospective feasibility study in polyneuropathy and Charcot Marie Tooth disease. Journal of Magnetic Resonance Imaging 2016;44:1513–1521.

17. Vaeggemose M, Vaeth S, Pham M, et al. Magnetic resonance neurography and diffusion tensor imaging of the peripheral nerves in patients with C harcot M arie T ooth Type 1A. Muscle & nerve 2017;56:E78–E84.

18. Kronlage M, Pitarokoili K, Schwarz D, et al. Diffusion tensor imaging in chronic inflammatory demyelinating polyneuropathy: diagnostic accuracy and correlation with electrophysiology. Investigative radiology 2017;52:701–707.

19. Chen Y, Baraz J, Xuan SY, et al. Multliparametric Quantitative MRI of Peripheral Nerves in the Leg: A Reliability Study. Journal of Magnetic Resonance Imaging 2024;59:563–574.

20. Li J, Krajewski K, Shy ME, Lewis RA. Hereditary neuropathy with liability to pressure palsy: the electrophysiology fits the name. Neurology 2002;58:1769–1773.

21. Fridman V, Sillau S, Bockhorst J, et al. Disease progression in charcot–marie–tooth disease related to MPZ mutations: A longitudinal study. Annals of neurology 2023;93:563–576.

22. Bai Y, Zhang X, Katona I, et al. Conduction block in PMP22 deficiency. Journal of Neuroscience 2010;30:600–608.

23. Shy M, Blake J, Krajewski K, et al. Reliability and validity of the CMT neuropathy score as a measure of disability. Neurology 2005;64:1209–1214.

24. Murphy SM, Herrmann DN, McDermott MP, et al. Reliability of the CMT neuropathy score (second version) in Charcot Marie Tooth disease. Journal of the Peripheral Nervous System 2011;16:191–198.

25. Chen Y, Moiseev D, Kong WY, Bezanovski A, Li J. Automation of Quantifying Axonal Loss in Patients with Peripheral Neuropathies through Deep Learning Derived Muscle Fat Fraction. Journal of Magnetic Resonance Imaging 2021;53:1539–1549.

26. Saba S, Chen Y, Maddipati KR, Hackett M, Hu B, Li J. Demyelination in hereditary sensory neuropathy type 1C. Annals of Clinical and Translational Neurology 2020;7:1502–1512.

27. Yeh FC, Liu L, Hitchens TK, Wu YL. Mapping immune cell infiltration using restricted diffusion MRI. Magnetic resonance in medicine 2017;77:603–612.

28. Pedregosa F, Varoquaux G, Gramfort A, et al. Scikit-learn: Machine learning in Python. the Journal of machine Learning research 2011;12:2825–2830.

29. Harris CR, Millman KJ, Van Der Walt SJ, et al. Array programming with NumPy. nature 2020;585:357–362.

30. Gommers R, Virtanen P, Haberland M, et al. scipy/scipy: SciPy 1.15. 0. Zenodo 2024.

31. García Andreu A, Costa M, Pastor O. Feature Selection in Medical Imaging: A Comprehensive Review. International Conference on Advanced Information Systems Engineering; 2025: Springer: 142–154.

32. Vijithananda SM, Jayatilake ML, Hewavithana B, et al. Feature extraction from MRI ADC images for brain tumor classification using machine learning techniques. Biomedical engineering online 2022;21:52.

33. Chen Y, Baraz J, Xuan SY, et al. Multiparametric quantitative MRI of peripheral nerves to differentiate axonal from demyelinating neuropathies (P11-8.002). Neurology 2023;100:2196.

34. Kollmer J, Kästel T, Jende JM, Bendszus M, Heiland S. Magnetization transfer ratio in peripheral nerve tissue: does it depend on age or location? Investigative Radiology 2018;53:397–402.

35. Kronlage M, Schwehr V, Schwarz D, et al. Peripheral nerve diffusion tensor imaging (DTI): normal values and demographic determinants in a cohort of 60 healthy individuals. European radiology 2018;28:1801–1808.

