## Supplemental Material for "Multiparametric Quantitative MRI of Peripheral Nerves to Differentiate Demyelinating from Axonal Polyneuropathies"

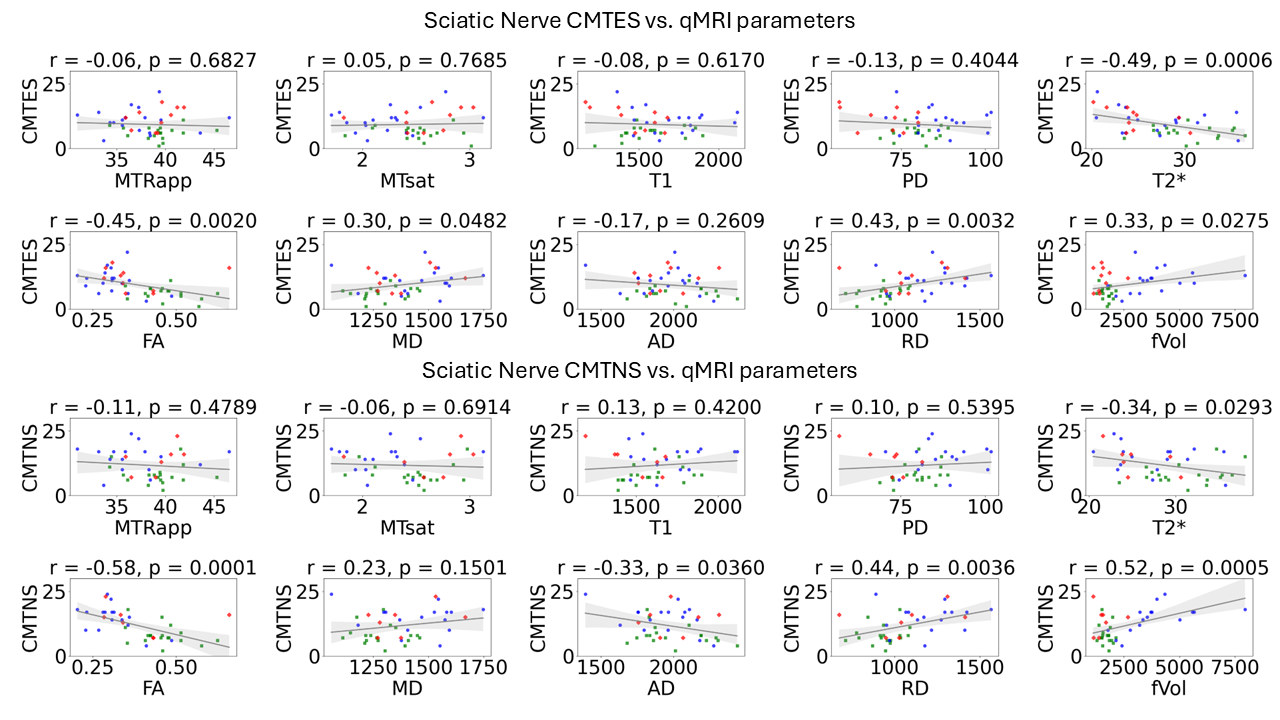

**Figure S1**. Scatter plots of examination (CMTES) and neuropathy (CMTNS) scores vs. qMRI parameters in the sciatic nerve. The result of a linear regression (gray line) is shown and in light gray the 5%-95% confidence interval of the regression. Green squares correspond to HNPP patients, Blue dots correspond to CMT1 patients, and red diamonds to CMT2 patients. In the upper part, the correlation coefficient (r) and p-values are shown for each plot.

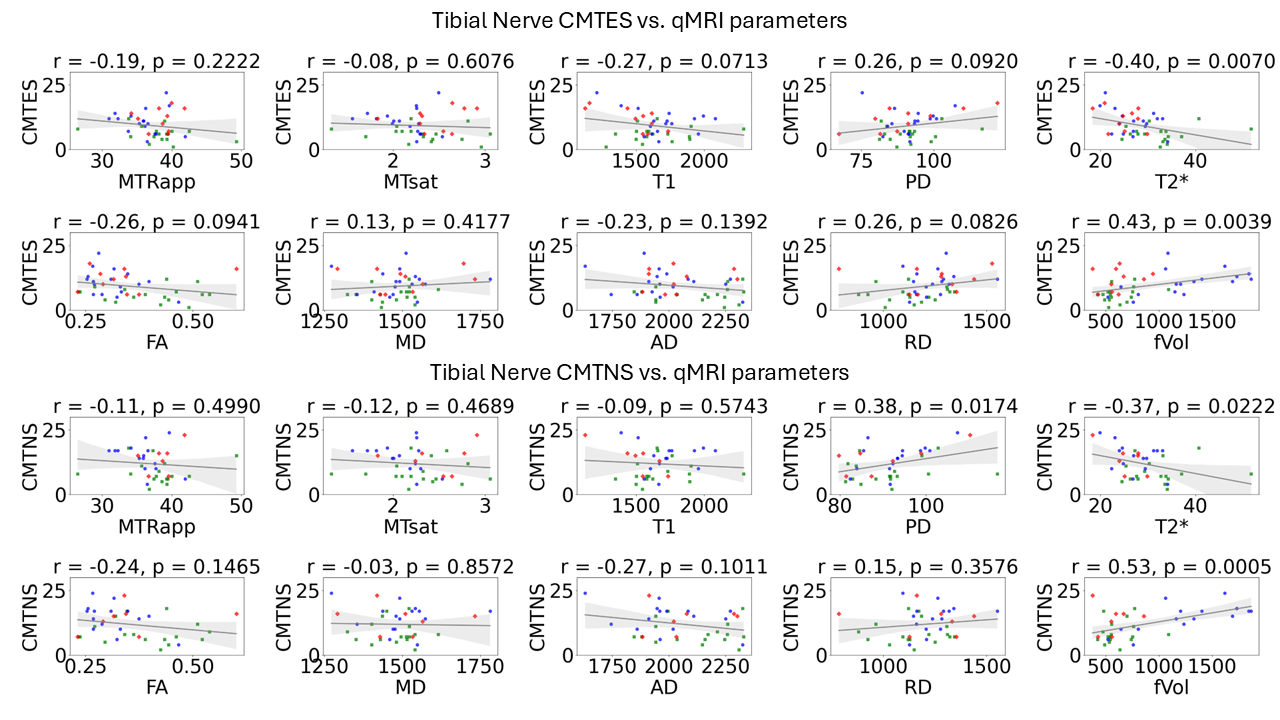

**Figure S2**. Scatter plots of examination (CMTES) and neuropathy (CMTNS) scores vs. qMRI parameters in the tibial nerve. The result of a linear regression (gray line) is shown and in light gray the 5%-95% confidence interval of the regression. Green squares correspond to HNPP patients, Blue dots correspond to CMT1 patients, and red diamonds to CMT2 patients. In the upper part, the correlation coefficient (r) and p-values are shown for each plot.

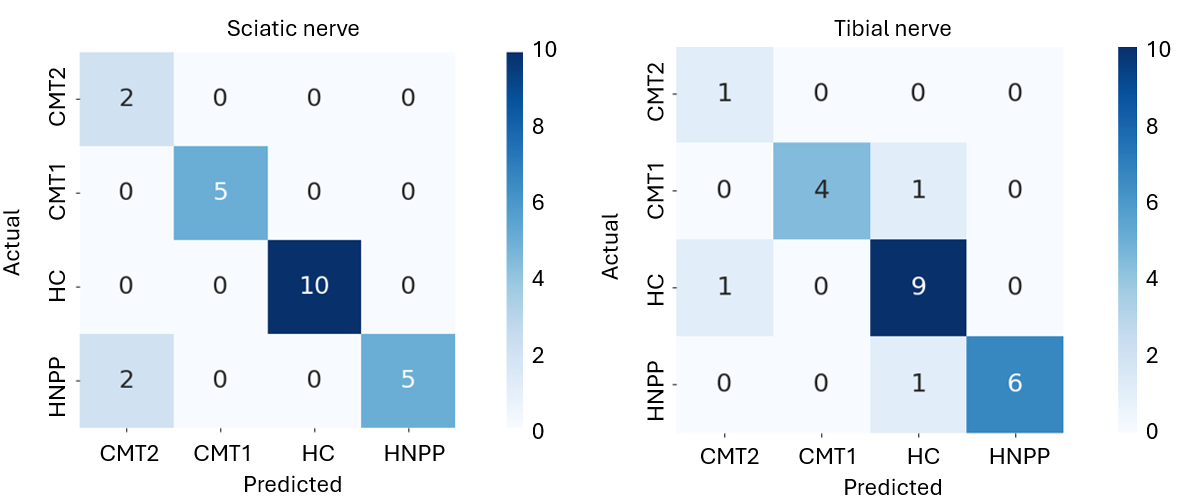

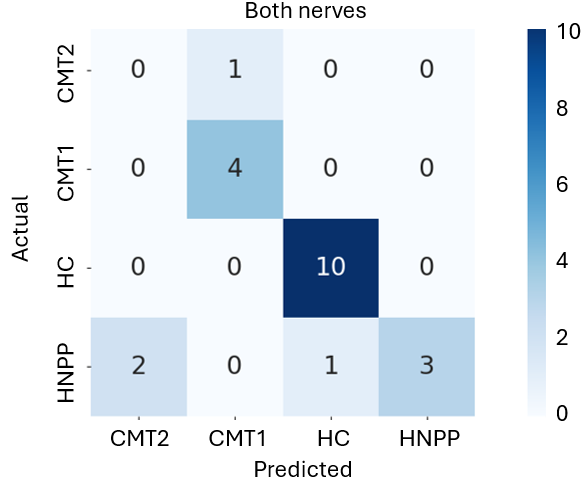

**Figure S3**. Confusion matrix (actual vs. predicted phenotype) of a test set (not seen during the model training) evaluated with the Logistic regression model from MP qMRI in the Sciatic nerve (left plot, n=24), the Tibial nerve (center plot, n=23) and both nerves (right plot, n=22). A blue color bar represents the number of patients in each category. Probabilities for all subjects used for this experiment are reported in Table S5.

**Table S1. Multiparametric qMRI protocol at 3T**

| **Sequence** | UHR  (3D GRE) | mDixon  (3D GRE) | MT_OFF  (3D GRE) | MT_ON  (3D GRE) | T1W  (3D GRE) | PDW  (3D GRE) | B1map  (3D GRE) | DTI  (2D SS-EPI) |
| --- | --- | --- | --- | --- | --- | --- | --- | --- |
| **TR (ms)** | 26 | 20 | 40 | 40 | 20 | 20 | 12 | 4000 |
| **# of echoes** | 1 | 6 | 6 | 3 | 2 | 2 | 1 | - |
| **1^st^ TE/ΔTE (ms)** | 5.1 | 2.1/3.0 | 5.1/5.24 | 5.1/5.24 | 5.1/5.24 | 5.1/5.24 | 5.1 | 93 |
| **FA (degree)** | 12 | 6 | 10 | 10 | 22 | 5 | 4, 17 |  |
| **FOV (mm^2^)** | 160×160 | 160×160 | 160×160 | 160×160 | 160×160 | 160×160 | 160×160 | 160×160 |
| **TH (mm)** | 3 | 3 | 3 | 3 | 3 | 3 | 3 | 6 |
| **Matrix (Read × Phase)** | 1024×768 | 128×128 | 256×192 | 256×192 | 256×192 | 256×192 | 256×256 | 128×128 |
| **Voxel Size (mm^3^)** | 0.15×0.15×3.0 | 1.2×1.2×3.0 | 0.6×0.6×3.0 | 0.6×0.6×3.0 | 0.6×0.6×3.0 | 0.6×0.6×3.0 | 0.6×0.6×3.0 | 1.2×1.2×6.0 |
| **# Of slices** | 40 | 40 | 40 | 40 | 40 | 40 | 40 | 16 |
| **Fat Suppression** | WE | None | WE | WE | WE | WE | None | SPAIR |
| **GRAPPA Factor** | 2 | 2 | 2 | 2 | 2 | 2 | 2 | 3 |
| **NEX** | 1 | 1 | 1 | 1 | 1 | 1 | 1 | 6 |
| **Other Parameters** | FC=slice | Readout Mode  = monopolar |  | MT=ON |  |  | FC= slice/  read/phase | b=800 s/mm^2^  20 directions |
| **TA (min:sec)** | 7:36 | 1:08 | 3:10 | 3:10 | 1:36 | 1:36 | 2:36 | 8:46 |

Abbreviations: UHR=ultra-high resolution; GRE=gradient recalled echo; MT=magnetization transfer; T1W=T1 weighted; PDW=proton density weighted; SS-EPI=single-shot echo planner imaging; TR=relaxation time; TE=echo time; FA=flip angle; FOV=field of view; TH=slice thickness; WE=water excitation; SPAIR=spectral adiabatic inversion recovery; GRAPPA=generalized auto-calibrating partially parallel acquisitions; FC=flow compensation; NEX=number of excitations or averages; TA=acquisition time or scan time.

**Table S2.** ANOVA test results for the individual qMRI parameters of different polyneuropathies phenotypes (CMT1, CMT2, HNPP) and the control group.

|  | **Sciatic nerve** | | **Tibial nerve** | |
| --- | --- | --- | --- | --- |
| **Parameter** | **F-statistic** | **p-value** | **F-statistic** | **p-value** |
| MTR | 4.09 | 9.54E-02 | 9.03 | 3.83E-04 |
| MTsat | 19.52 | 2.31E-08 | 16.27 | 3.78E-07 |
| T1 | 20.18 | 1.36E-08 | 16.40 | 4.54E-07 |
| PD | 19.13 | 3.18E-08 | 5.26 | 2.47E-02 |
| T2* | 6.59 | 5.13E-03 | 4.56 | 5.53E-02 |
| FA | 32.34 | 1.41E-12 | 26.52 | 1.43E-10 |
| MD | 6.54 | 5.39E-03 | 5.09 | 2.98E-02 |
| AD | 6.83 | 3.90E-03 | 8.72 | 5.34E-04 |
| RD | 23.19 | 9.27E-10 | 18.79 | 4.48E-08 |
| fVol | 40.99 | 7.30E-15 | 39.99 | 2.53E-14 |

**Table S3.** CMTES and CMTNS qMRI score equation coefficients and their relative weights for the sciatic and tibial nerves.

|  | **Sciatic Nerve** | | | | **Tibial Nerve** | | | | **Combined Nerves** | | | |
| --- | --- | --- | --- | --- | --- | --- | --- | --- | --- | --- | --- | --- |
|  | **CMTES** |  | **CMTNS** |  | **CMTES** |  | **CMTNS** |  | **CMTES (Tn./Sn.)** | | **CMTNS (Tn./Sn.)** | |
| **qMRI param.** | **(min / max =**  **-28.65 / -13.65)** | **Var. (%)** | **(min / max =**  **-34.55 / -19.76)** | **Var. (%)** | **(min / max = -69.73 / -55.03)** | **Var. (%)** | **(min / max = --52.84 / -31.24)** | **Var. (%)** | **(min / max = -33.03 / -17.99)** | **Var. (%)** | **(min / max = -12.04 / 18.99)** | **Var. (%)** |
| MTR | -0.1338 | 0.49 | -0.1973 | 0.79 | -0.9480 | 7.00 | -0.6659 | 4.89 | -0.41 / -0.21 | 2.37 / 1.14 | 0.17 / -0.52 | 0.36 / 0.84 |
| Mtsat | 0.9060 | 0.41 | 6.3029 | 2.88 | -4.2709 | 3.78 | -1.7723 | 1.29 | -12.22 / 10.79 | 8.37 / 7.02 | 0.67 / 5.23 | 0.13 / 1.03 |
| T1 | -0.0034 | 0.92 | -0.0030 | 0.74 | -0.0130 | 7.29 | -0.0078 | 3.33 | -0.01 / 0.01 | 5.58 / 4.96 | -0.02 / 0.01 | 2.11 / 1.08 |
| PD | 0.0566 | 0.71 | 0.1734 | 1.97 | 0.0781 | 1.89 | 0.3188 | 5.28 | 0.13 / 0.01 | 2.41 / 0.25 | 0.17 / 0.32 | 0.75 / 1.62 |
| T2* | -0.5264 | 2.91 | -0.2016 | 1.14 | -0.5606 | 7.70 | -0.7175 | 8.50 | -0.58 / -0.39 | 6.10 / 3.09 | 0.31 / -0.94 | 1.00 / 2.25 |
| FA | -28.3508 | 3.73 | -94.7853 | 12.24 | 34.4789 | 7.49 | -37.16 | 6.75 | 24.18 / -21.92 | 4.06 / 4.24 | 54.49 / -26.89 | 2.72 / 1.48 |
| MD | 0.2070 | 39.30 | 0.1703 | 33.47 | -0.1019 | 24.73 | -0.1411 | 27.54 | -0.04 / -0.04 | 7.94 / 11.77 | -0.14 / 0.35 | 7.36 / 28.95 |
| AD | -0.0627 | 16.04 | -0.0429 | 11.47 | 0.0257 | 10.48 | 0.0596 | 20.14 | 0.00 / 0.03 | 2.50 / 11.10 | 0.07 / -0.14 | 6.47 / 15.77 |
| RD | -0.1424 | 35.50 | -0.1417 | 35.31 | 0.0809 | 29.64 | 0.0753 | 22.29 | 0.03 / 0.02 | 9.30 / 7.80 | 0.06 / -0.20 | 4.97 / 21.11 |

**Table S4.** ROC AUC values from MP qMRI Logistic Regression model for different individual parameters and parameters combinations.

|  | **CMT1** | | | **CMT2** | | | **HNPP** | | | **HC** | | | | |
| --- | --- | --- | --- | --- | --- | --- | --- | --- | --- | --- | --- | --- | --- | --- |
| Parameters | **AUC sciatic n.** | **AUC tibial n.** | **AUC combined** | **AUC sciatic n.** | **AUC tibial n.** | **AUC combined** | **AUC sciatic n.** | **AUC tibial n.** | **AUC combined** | **AUC sciatic n.** | **AUC tibial n.** | **AUC combined** | **Average** | **Std** |
| MTR | 0.77 | 0.76 | 0.74 | 0.57 | 0.55 | 0.49 | 0.55 | 0.45 | 0.55 | 0.78 | 0.68 | 0.76 | 0.64 | 0.12 |
| MTsat | 0.79 | 0.84 | 0.82 | 0.79 | 0.76 | 0.76 | 0.59 | 0.66 | 0.65 | 0.84 | 0.84 | 0.85 | 0.77 | 0.08 |
| T1 | 0.76 | 0.80 | 0.78 | 0.79 | 0.54 | 0.71 | 0.63 | 0.64 | 0.65 | 0.92 | 0.86 | 0.91 | 0.75 | 0.11 |
| PD | 0.67 | 0.78 | 0.76 | 0.39 | 0.79 | 0.76 | 0.62 | 0.67 | 0.73 | 0.62 | 0.67 | 0.73 | 0.68 | 0.10 |
| T2* | 0.64 | 0.64 | 0.59 | 0.67 | 0.76 | 0.74 | 0.68 | 0.71 | 0.70 | 0.52 | 0.50 | 0.52 | 0.64 | 0.08 |
| FA | 0.85 | 0.89 | 0.88 | 0.65 | 0.71 | 0.70 | 0.55 | 0.79 | 0.74 | 0.89 | 0.88 | 0.90 | 0.79 | 0.11 |
| MD | 0.69 | 0.75 | 0.74 | 0.43 | 0.55 | 0.38 | 0.56 | 0.63 | 0.66 | 0.75 | 0.63 | 0.78 | 0.63 | 0.12 |
| AD | 0.76 | 0.67 | 0.73 | 0.64 | 0.63 | 0.66 | 0.38 | 0.46 | 0.37 | 0.77 | 0.77 | 0.78 | 0.64 | 0.14 |
| RD | 0.83 | 0.87 | 0.87 | 0.53 | 0.65 | 0.68 | 0.54 | 0.71 | 0.73 | 0.90 | 0.82 | 0.90 | 0.75 | 0.13 |
| fVol | 0.92 | 0.97 | 0.94 | 0.57 | 0.62 | 0.64 | 0.71 | 0.62 | 0.68 | 0.63 | 0.67 | 0.63 | 0.72 | 0.14 |
| Comb 1 | 0.95 | 0.89 | 0.95 | 0.85 | 0.73 | 0.79 | 0.82 | 0.87 | 0.88 | 0.88 | 0.90 | 0.94 | 0.87 | 0.06 |
| Comb 2 | 0.95 | 0.94 | 0.94 | 0.80 | 0.66 | 0.75 | 0.85 | 0.83 | 0.85 | 0.87 | 0.91 | 0.92 | 0.86 | 0.08 |
| All | 0.94 | 0.97 | 0.90 | 0.86 | 0.72 | 0.83 | 0.82 | 0.88 | 0.85 | 0.88 | 0.90 | 0.94 | 0.87 | 0.06 |

**Table S5.** Assigned probabilities from the MP qMRI Logistic Regression model evaluation in test set for individual subjects.

|  |  | **Sciatic Nerve** | |  |  | **Tibial Nerve** | |  |  | **Both nerves** | |  |  |
| --- | --- | --- | --- | --- | --- | --- | --- | --- | --- | --- | --- | --- | --- |
| **Sub. Idx.** | **Phenotype** | **CMT2 Prob.** | **CMT1 Prob** | **HC Prob** | **HNPP Prob** | **CMT2 Prob.** | **CMT1 Prob** | **HC Prob** | **HNPP Prob** | **CMT2 Prob.** | **CMT1 Prob** | **HC Prob** | **HNPP Prob** |
| 1 |  |  |  |  |  | 0.0643 | 0.0431 | 0.2209 | **0.6717** |  |  |  |  |
| 2 | HNPP | 0.0849 | 0.0020 | 0.0258 | **0.8873** | 0.1392 | 0.0067 | 0.0159 | **0.8382** | **0.9984** | 0.0000 | 0.0010 | 0.0006 |
| 3 | HNPP | **0.8194** | 0.0047 | 0.0262 | 0.1497 | 0.2284 | 0.0087 | **0.6995** | 0.0635 | 0.0000 | 0.0000 | 0.0000 | **1.0000** |
| 4 | HNPP | 0.0153 | 0.0136 | 0.0102 | **0.9609** | 0.0527 | 0.0017 | 0.0070 | **0.9385** | 0.0000 | 0.0005 | 0.4490 | **0.5505** |
| 5 | HNPP | 0.0756 | 0.0065 | 0.0042 | **0.9137** |  |  |  |  |  |  |  |  |
| 6 | HNPP | 0.0028 | 0.0061 | 0.0209 | **0.9701** | 0.0105 | 0.0112 | 0.0477 | **0.9306** | **0.7857** | 0.0159 | 0.0033 | 0.1951 |
| 7 | HNPP | **0.5454** | 0.0464 | 0.0075 | 0.4006 | 0.0011 | 0.0088 | 0.0220 | **0.9681** | 0.3422 | 0.0310 | **0.6239** | 0.0028 |
| 8 | HNPP | 0.0875 | 0.0250 | 0.1441 | **0.7434** | 0.0009 | 0.0090 | 0.0337 | **0.9565** | 0.0008 | 0.0000 | 0.0000 | **0.9992** |
| 9 | CMT1B | 0.0068 | **0.7893** | 0.0006 | 0.2033 | 0.0538 | 0.1827 | **0.4787** | 0.2848 | 0.0000 | **0.9999** | 0.0001 | 0.0000 |
| 10 | CMT1A | 0.0000 | **1.0000** | 0.0000 | 0.0000 | 0.0000 | **0.9935** | 0.0066 | 0.0006 | 0.0000 | **1.0000** | 0.0000 | 0.0000 |
| 11 | CMT1A | 0.0000 | **1.0000** | 0.0000 | 0.0000 | 0.0000 | **0.9725** | 0.0275 | 0.0000 | 0.0000 | **1.0000** | 0.0000 | 0.0000 |
| 12 | CMT1A | 0.0000 | **1.0000** | 0.0000 | 0.0000 | 0.0000 | **0.9935** | 0.0064 | 0.0000 | 0.0000 | **1.0000** | 0.0000 | 0.0000 |
| 13 | CMT1A | 0.0000 | **1.0000** | 0.0000 | 0.0000 | 0.0000 | **0.9627** | 0.0371 | 0.0002 | 0.0000 | **1.0000** | 0.0000 | 0.0000 |
| 14 | CMT2D | **0.9555** | 0.0003 | 0.0073 | 0.0370 |  |  |  |  |  |  |  |  |
| 15 | CMT2B | **0.6209** | 0.2187 | 0.0156 | 0.1448 | **0.6303** | 0.0365 | 0.0001 | 0.3331 | 0.0275 | **0.9688** | 0.0000 | 0.0037 |
| 16 | Control | 0.0360 | 0.0006 | **0.8958** | 0.0676 | 0.0138 | 0.0014 | **0.9782** | 0.0066 | 0.0037 | 0.0000 | **0.9963** | 0.0000 |
| 17 | Control | 0.0224 | 0.0006 | **0.8707** | 0.1063 | 0.0016 | 0.0004 | **0.9946** | 0.0034 | 0.0000 | 0.0000 | **1.0000** | 0.0000 |
| 18 | Control | 0.0013 | 0.0003 | **0.9117** | 0.0867 | 0.0010 | 0.0006 | **0.9387** | 0.0597 | 0.0000 | 0.0000 | **1.0000** | 0.0000 |
| 19 | Control | 0.0051 | 0.0029 | **0.9550** | 0.0371 | 0.0078 | 0.0029 | **0.9823** | 0.0070 | 0.0010 | 0.0000 | **0.9988** | 0.0002 |
| 20 | Control | 0.0003 | 0.0397 | **0.8318** | 0.1281 | 0.0001 | 0.0266 | **0.9257** | 0.0476 | 0.0000 | 0.0000 | **0.9997** | 0.0003 |
| 21 | Control | 0.1169 | 0.0015 | **0.8151** | 0.0666 | 0.0050 | 0.0019 | **0.9894** | 0.0037 | 0.0042 | 0.0000 | **0.9957** | 0.0001 |
| 22 | Control | 0.0334 | 0.0027 | **0.8859** | 0.0780 | 0.1069 | 0.0084 | **0.8400** | 0.0447 | 0.0143 | 0.0000 | **0.9834** | 0.0022 |
| 23 | Control | 0.1181 | 0.0092 | **0.7845** | 0.0881 | 0.1469 | 0.1654 | **0.6132** | 0.0744 | 0.2220 | 0.0004 | **0.7752** | 0.0024 |
| 24 | Control | 0.1362 | 0.0003 | **0.8034** | 0.0601 | 0.0246 | 0.0036 | **0.9481** | 0.0238 | 0.0012 | 0.0000 | **0.9988** | 0.0000 |
| 25 | Control | 0.0061 | 0.0008 | **0.9848** | 0.0082 | **0.4290** | 0.1195 | 0.3525 | 0.0990 | 0.0394 | 0.0002 | **0.9595** | 0.0009 |

***CMT Imaging Score (CMTIS) to Predict Disease Severity:***

To derive the CMTIS predictive of CMTNS and CMTES, a linear regression model was trained using Python Scikit-learn ^1^. Stratified 5-fold cross-validation was employed to preserve the distribution of polyneuropathy subgroups (CMT1, CMT2, HNPP) across folds. For each training fold, a linear regression model was fit using the equation:

(1) ${\hat{y}=\beta}_{0}+\sum_{i=1}^{n} \beta_{i}x_{i}$

where *ŷ* is the predicted CMTNS or CMTES, *xᵢ* are the *n* qMRI variables, and *βᵢ* are the learned coefficients. The average of the coefficients across folds was computed to obtain a stable estimate of each variable's contribution.

The unscaled CMTIS was subsequently computed for each subject as a weighted sum of the imaging variables:

(2) $Unscaled CMTIS=\sum_{i=1}^{n} \bar{\beta}_{i}x_{i}$

where *β̄ᵢ* denotes the average coefficient for the *i*-th qMRI parameter from cross-validation. This score was then linearly scaled to the range [0, 36] as the final CMTIS to match the CMTNS range (which contains the CMTES range [0 - 28]), using the observed minimum and maximum values:

(3) *CMTIS = (Unscaled CMTIS - min) / (max - min) × 36*

Pearson correlation between the CMTIS and CMTNS was computed using SciPy ^2^. The strength and significance of the correlation were reported as the Pearson correlation coefficient r and associated p-value.

Finally, to assess the relative importance of each qMRI variable, a variance decomposition was performed. The contribution of each variable was computed as:

(4) *Normalized Contributionᵢ* =$100*\frac{\left| \bar{\beta}_{i}\sigma_{i} \right|}{\sum_{j} \left| \bar{\beta}_{j}\sigma_{j} \right|}$ , 100% = Σ*_i_ Normalized Contribution_i_*

where *σᵢ* is the standard deviation of variable *xᵢ*. Contributions were normalized to sum to 100% to yield a percentage-based importance metric.

***References:***

1.	Pedregosa F, Varoquaux G, Gramfort A, et al. Scikit-learn: Machine learning in Python. the Journal of machine Learning research 2011;12:2825-2830.

2.	Gommers R, Virtanen P, Haberland M, et al. scipy/scipy: SciPy 1.15. 0. Zenodo 2024.
